# Four Movement Screen Structure (4MS): A Theoretical Framework for Understanding Postural Control Structures Underlying Activities of Daily Living and an Exploratory Cross-Sectional Study

**DOI:** 10.64898/2026.05.03.26352310

**Authors:** Hiroshi Osato

## Abstract

**Background:** Activities of Daily Living (ADL) assessments are essential outcome measures in rehabilitation and long-term care, but primarily focus on task completion and provide limited insight into the postural control structures underlying movement failure. This paper proposes the Four Movement Screen Structure (4MS), a theoretical framework that reconceptualizes human movement control through four postural control phases: supine, sitting, standing, and single-leg standing. The framework proposes that functional decline may present with non-continuity, asymmetry, and compensatory preservation, rather than a simple reversal of motor development.

**Methods:** An exploratory, hypothesis-generating cross-sectional study was conducted with 297 certified care recipients (mean age 80.5 years) across multiple day-service facilities in Japan. Each participant was assessed using both the Barthel Index (BI) and the 4MS evaluation. Descriptive statistics, Pearson correlations, chi-square tests, and Fisher’s exact tests were used to explore the structural properties of the framework.

**Results:** The mean BI total was 89.0 (SD = 13.8); the mean 4MS total score was 7.75 (SD = 2.02). A moderate positive correlation was found between BI total and 4MS total score (r = 0.471, p < 0.001, 95% CI [0.378, 0.555]). Of the five defined decline types, four were observed: mixed (57.6%), supine-dominant (21.2%), standing-dominant (5.7%), and single-leg-dominant (15.5%); sitting-dominant was not observed. The supine phase was the primary intervention target in 74.4% of cases—a finding we term the “supine paradox.” In a subsample of 274 participants, 90.0% of those in the low supine score group (0–1.0, n = 170) performed rising from supine independently, suggesting that this paradox reflects qualitative deficits in foundational motor control masked by compensatory strategies.

**Conclusions:** These exploratory findings are broadly consistent with the non-reversal hypothesis and suggest that 4MS may capture structural dimensions of postural control not fully represented by conventional ADL assessment. As a hypothesis-generating study, these findings should be interpreted as generating testable hypotheses for future longitudinal and interventional research. Keywords: Postural control; Activities of daily living; Motor development; Functional decline; Barthel Index; Long-term care; Supine paradox; Non-reversal hypothesis; Geriatric assessment; Exploratory study

## Background

Assessment of Activities of Daily Living (ADL) plays a central role in rehabilitation medicine and long-term care practice. Established instruments such as the Barthel Index (BI) [1] and the Functional Independence Measure (FIM) [2, 3] have been widely used to quantify functional independence and to support clinical decision-making and policy design.

However, these assessments primarily describe the outcome of movement performance. They provide limited insight into why a particular activity cannot be performed, and they do not offer a theoretical framework for explaining the underlying postural or motor control structures that constrain movement execution.

In clinical practice, it is not uncommon to observe individuals who are able to perform certain ADL tasks while experiencing marked difficulty with others. Such inconsistencies are difficult to explain solely through task-based evaluation and suggest the need for a theoretical framework that focuses on the postural control structures underlying movement.

To address this gap, this paper proposes the Four Movement Screen Structure (4MS). The 4MS reconceptualizes ADL not as isolated tasks but as movements performed under four postural control phases—supine, sitting, standing, and single-leg standing. This framework shares theoretical foundations with developmental kinesiology [4, 5] and the Dynamic Neuromuscular Stabilization (DNS) approach [6, 7], applying the developmental sequence of postural control acquisition to adult functional assessment. The framework draws on classical descriptions of motor development [8, 9, 10, 11, 12, 13] and theories of postural control [14, 15, 16, 17].

As a structural hypothesis, 4MS proposes that functional decline does not follow a simple reversal of developmental sequence (non-reversal hypothesis), but may exhibit three characteristics: (1) non-continuity—deficits appearing in non-adjacent phases, (2) asymmetry—side-to-side differences within a single phase, and (3) compensatory preservation—higher phases maintained despite lower-phase deficits. This hypothesis provides an alternative perspective to Reisberg et al.’s [18] retrogenesis theory. The 4MS does not claim to replace existing motor control theories [19] or established ADL instruments, but rather complements them by addressing the structural dimension of postural control conditions.

The objective of this study is to present the 4MS framework and to describe, using data from 297 certified care recipients, the extent to which cross-sectional patterns are consistent with the proposed framework, thereby generating testable hypotheses for future confirmatory research. This study is positioned as an exploratory, hypothesis-generating investigation.

## Methods

To examine the structural properties proposed by the 4MS framework, a cross-sectional observational study was conducted. This study is exploratory and hypothesis-generating rather than confirmatory. Reporting follows the STROBE (Strengthening the Reporting of Observational Studies in Epidemiology) statement for cross-sectional studies.

### Participants

A total of 301 certified care recipients were enrolled across multiple day-service (通所介護) facilities in Aomori Prefecture, Japan. After excluding 4 cases with incomplete data matching, 297 participants were included in the final analysis.

**Table 1.** Participant Characteristics (N = 297)

| Variable | Value |
| --- | --- |
| Age, years, mean | 80.5 (8.7) |
| Age range | 52–101 |
| Male, n (%) | 117 (39.4%) |
| Female, n (%) | 180 (60.6%) |
| Support Level 1, n (%) | 1 (0.3%) |
| Support Level 2, n (%) | 1 (0.3%) |
| Care Level 1, n (%) | 139 (46.8%) |
| Care Level 2, n (%) | 116 (39.1%) |
| Care Level 3, n (%) | 28 (9.4%) |
| Care Level 4, n (%) | 9 (3.0%) |
| Care Level 5, n (%) | 3 (1.0%) |

### Assessments

Barthel Index (BI): The standard 10-item BI was administered by trained functional training instructors and care staff [1, 20, 21]. Total score range: 0–100.

4MS Evaluation: Each participant was assessed using the 4MS framework. The 4MS evaluation employs a sequential Yes/No screening procedure for each of the four postural control phases.

Within each phase, the examiner observes whether the participant can perform progressively demanding movement tasks. Scoring proceeds sequentially: if the participant cannot perform the first (lowest-level) task, the score is 0; if the first task is achieved but the next is not, the score is 1; and so on up to a maximum of 3. The supine-phase screening items are ordered according to the developmental motor sequence. “Pelvic stability” in the supine phase is assessed as an observable indicator of sagittal-plane (anterior-posterior) pelvic tilt control. This corresponds to the approximately 3-month sagittal stabilization milestone and represents the clinical manifestation of deep stabilizer function achieved through intra-abdominal pressure (IAP) generated by coordinated activation of the diaphragm, pelvic floor muscles, transversus abdominis, and multifidus [4, 5, 6, 7]. In developmental kinesiology, the neonatal trunk (0 months) is characterized by anterior pelvic tilt and rib flare, reflecting the absence of IAP; by approximately 3 months, the establishment of opposing diaphragm–pelvic floor activation generates IAP, achieving sagittal pelvic stabilization. From a developmental motor science perspective, the first cervical control emerges as cervical extension in prone (approximately 1–2 months), while active cervical flexion in supine is a later achievement (4–5 months).

Accordingly, the screening items correspond to the supine developmental sequence: respiratory stabilization and cervical stability (Score 1) → sagittal plane pelvic stabilization and hip flexion (Score 2) → active cervical flexion with trunk rotation (Score 3: rolling). Cervical intervention is addressed in a separate intervention protocol. The specific scoring criteria for each phase are presented in Table 1a.

**Table 1a.**
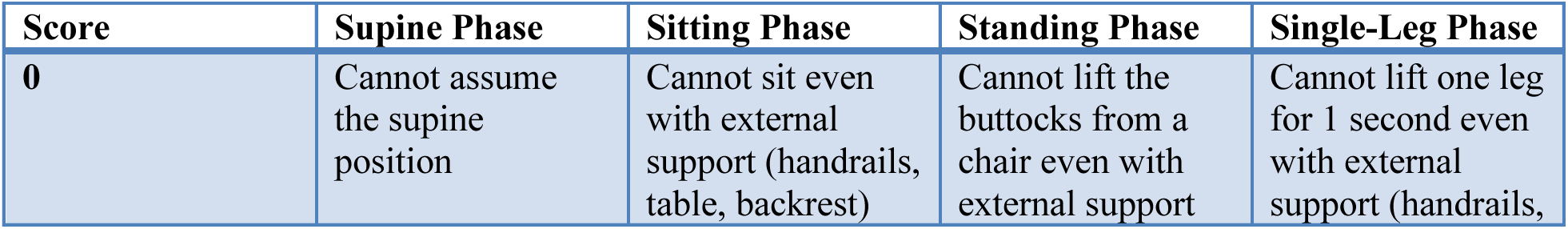
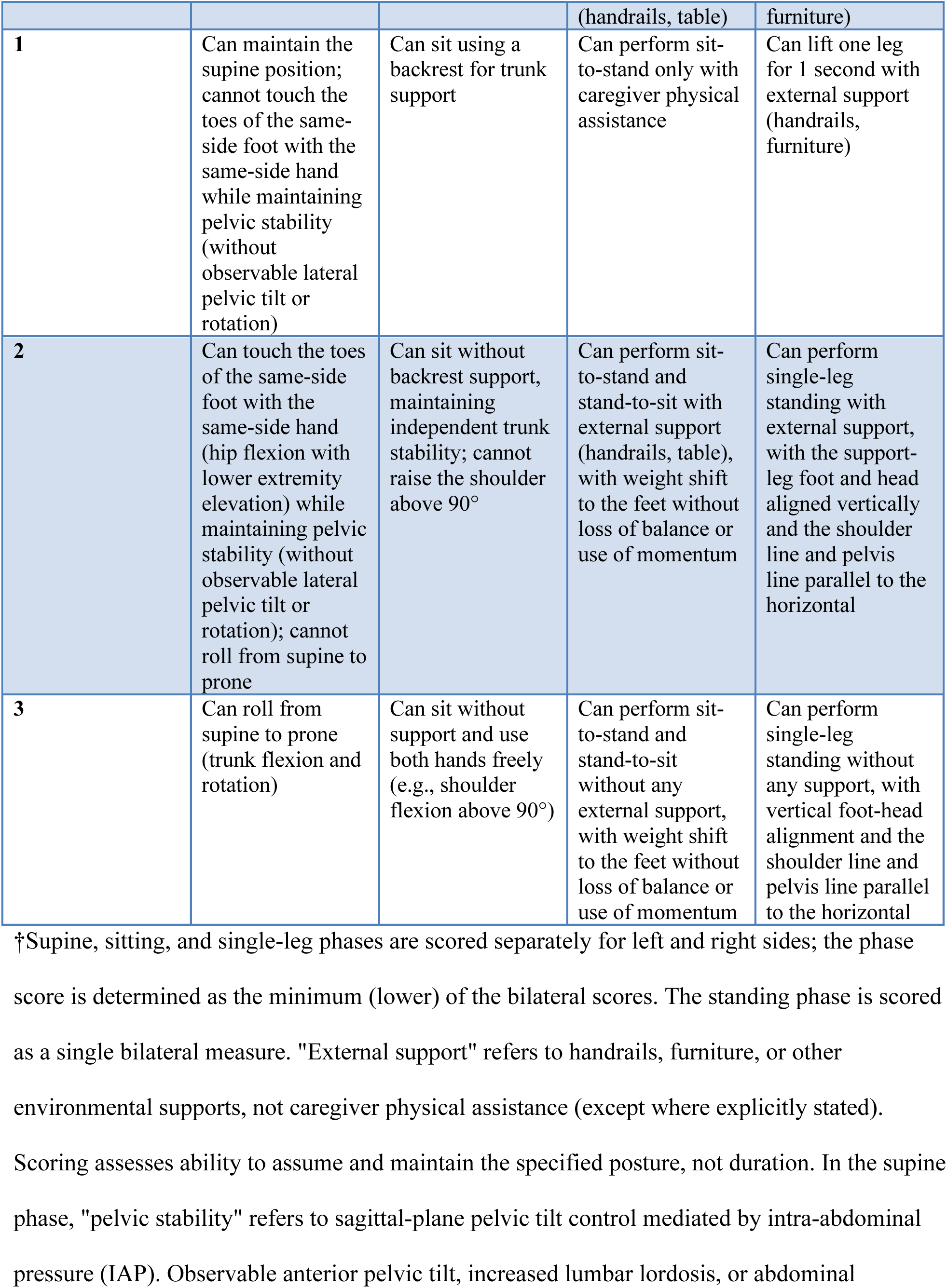
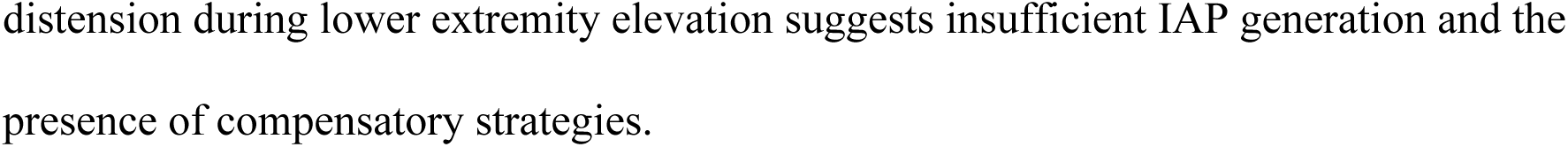
4MS Scoring Criteria for Each Postural Control Phase†.

The 4MS total score was calculated as the sum of the four phase scores (range: 0–12). Left-right difference (|left − right|) was recorded as a measure of asymmetry for each applicable phase.

BI Phase Reclassification: To examine the structural relationship between BI and 4MS, each BI item was reclassified into the postural phase most closely associated with its primary postural control requirement:

| BI Item | Phase Classification |
| --- | --- |
| Feeding, Grooming, Dressing, Bowel, Bladder | Sitting |
| Transfer, Toilet | Standing |
| Bathing, Walking, Stairs | Single-Leg |

This BI item reclassification is based on the primary postural control condition required for each ADL task and does not imply that the 4MS directly assesses that ADL task. For example, walking is classified under the single-leg phase because it requires single-leg postural control, but the 4MS single-leg phase scoring criteria evaluate single-leg standing ability per se.

### Analysis Variables

Decline type classification: Based on the 4MS scores across four phases, each participant was classified into one of five decline types. The classification rule was as follows: (a) if a single phase had the uniquely lowest score among the four phases, the decline was classified as that phase-dominant type (supine-dominant, sitting-dominant, standing-dominant, or single-leg-dominant); (b) if two or more phases shared the lowest score, or if no single phase showed a clear minimum, the decline was classified as mixed type. This classification identifies the primary structural pattern of postural control deficit for each individual. This decline type classification was conducted as a pre-defined, rule-based classification derived from the 4MS theoretical framework, and no exploratory clustering methods were employed.

Target phase: The postural phase identified as the primary intervention target, determined by the 4MS evaluation algorithm. The algorithm applies the following sequential rules: (1) if two or more phases share the same score, the developmental order (supine → sitting → standing → single-leg) is followed, prioritizing the developmentally earlier (foundational) phase; (2) if left-right asymmetry exists within a phase, the lower of the bilateral scores determines that phase’s priority ranking. This algorithm produces a single target phase per individual, representing the postural control phase with the greatest relative deficit weighted toward foundational (developmentally earlier) phases.

ADL cause phase: The postural phase most closely associated with the participant’s primary ADL limitations, determined through clinical assessment. When a participant exhibited difficulty in specific ADL tasks, the task was mapped to its corresponding postural control phase based on the BI phase reclassification (Table 1), and the phase accounting for the greatest ADL limitation was identified as the ADL cause phase. Participants without identifiable ADL difficulty were classified as “no identified difficulty.” Notably, the Barthel Index does not include any item directly corresponding to supine-phase postural control; therefore, the supine phase does not appear as an ADL cause phase in this classification.

### Statistical Analysis

Descriptive statistics were calculated for all variables. The normality of distributions was assessed using the Shapiro-Wilk test. Pearson correlation coefficients with 95% confidence intervals (Fisher z-transformation) were computed to examine associations between BI and 4MS scores. As a sensitivity analysis to account for the non-normal distribution of BI scores, Spearman rank correlation coefficients were also calculated. Frequency distributions were calculated for decline types and target phases. All analyses were performed using Python 3.11 with SciPy 1.11.

All analyses in this study are exploratory and descriptive in nature and are not intended as formal hypothesis tests. Accordingly, reported p-values should be interpreted descriptively, and interpretation is based primarily on effect sizes and 95% confidence intervals rather than on statistical significance thresholds. No a priori sample size calculation or formal power analysis was conducted, and no formal correction for multiple comparisons was applied, consistent with the hypothesis-generating purpose of the study.

## Results

### Barthel Index and 4MS Scores

The mean BI total score was 89.0 (SD = 13.8, range: 20–100).

**Table 2.**
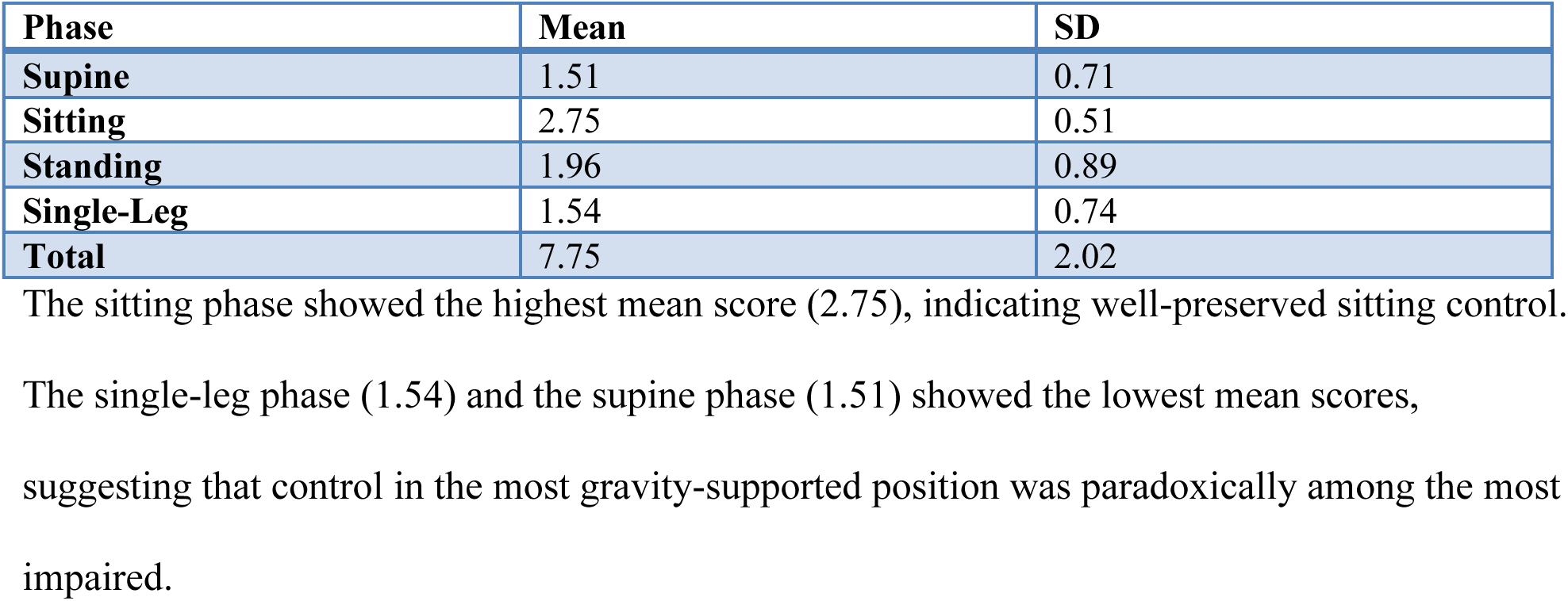
4MS Postural Control Scores (N = 297)

### Correlation Between BI and 4MS Scores

**Table 3.** Pearson Correlations Between BI Total and 4MS Phase Scores (with p-values and 95% CIs)

| Variable | r | p | 95% CI | N |
| --- | --- | --- | --- | --- |
| 4MS Total Score | 0.471 | < 0.001 | [0.378, 0.555] | 297 |
| Supine Score | 0.261 | < 0.001 | [0.152, 0.364] | 297 |
| Sitting Score | 0.238 | < 0.001 | [0.128, 0.343] | 297 |
| Standing Score | 0.375 | < 0.001 | [0.273, 0.469] | 297 |
| Single-Leg Score | 0.418 | < 0.001 | [0.319, 0.507] | 297 |

### Table 3a. Spearman Rank Correlations (Sensitivity Analysis)

Spearman rank correlations were consistent with Pearson results: 4MS Total (ρ = 0.451, p < 0.001). The moderate attenuation in Spearman coefficients relative to Pearson values is consistent with the ceiling effect in BI scores (36.0% at maximum score of 100), which compresses rank-based variation.

### Bilateral Scores and Asymmetry

Bilateral scores were available for supine, sitting, and single-leg phases (N = 297). Asymmetry (any left-right difference > 0) was present in 32.7% of participants (97/297). Phase-specific asymmetry prevalence was: supine 19.9% (59/297), sitting 12.5% (37/297), and single-leg 13.1% (39/297). Wilcoxon signed-rank tests revealed no systematic laterality bias in any phase (supine W = 876, p = 0.942; sitting W = 285, p = 0.250; single-leg W = 361, p = 0.650). Mean absolute left-right differences were: supine 0.242 (SD = 0.522), sitting 0.125 (SD = 0.331), and single-leg 0.138 (SD = 0.365).

### Decline Type Distribution

**Table 4.** Decline Type Distribution.

| Decline Type | N | % |
| --- | --- | --- |
| Mixed | 171 | 57.6% |
| Supine-dominant | 63 | 21.2% |
| Standing-dominant | 17 | 5.7% |
| Single-leg-dominant | 46 | 15.5% |
| Sitting-dominant | 0 | 0.0% |

Of the five decline types, four were observed. Sitting-dominant type did not appear in this sample, which is consistent with the generally high sitting phase scores (mean 2.75); no participant had the sitting phase as the uniquely lowest score. No single pattern dominated exclusively, and 42.4% of participants showed a single-phase-dominant pattern, consistent with structural heterogeneity in functional decline.

### Target Phase Distribution

**Table 5.** Target Phase Distribution.

| Target Phase | N | % |
| --- | --- | --- |
| Supine | 221 | 74.4% |
| Standing | 43 | 14.5% |
| Single-Leg | 32 | 10.8% |
| Sitting | 1 | 0.3% |

The supine phase was identified as the primary intervention target in 74.4% of cases.

### ADL Cause Phase Distribution

| ADL Cause Phase | N | % |
| --- | --- | --- |
| No identified difficulty | 112 | 41.2% |
| Single-leg phase | 107 | 39.3% |
| Sitting phase | 26 | 9.6% |
| Mixed | 26 | 9.6% |
| Standing phase | 1 | 0.4% |

Note: ADL cause phase was determinable for 272 of 297 participants; 25 cases lacked sufficient clinical information for phase attribution.
When ADL difficulty was present, single-leg phase limitations were the most frequent source (39.3%).

### Supine Score and Bed Mobility Independence

To quantitatively examine the supine paradox, the association between 4MS supine scores and bed mobility independence (rising from supine and rolling) was analyzed. Of the full sample of 297 participants, a subsample of 274 with available routine clinical records documenting basic movement (rising from supine and rolling) performance levels was included in this analysis. Performance levels were recorded on a four-level scale (independent, supervision, partial assistance, full assistance) and matched with 4MS supine scores (subsample mean supine score = 1.49, SD = 0.70, comparable to the full sample mean of 1.51).

**Table 6.**
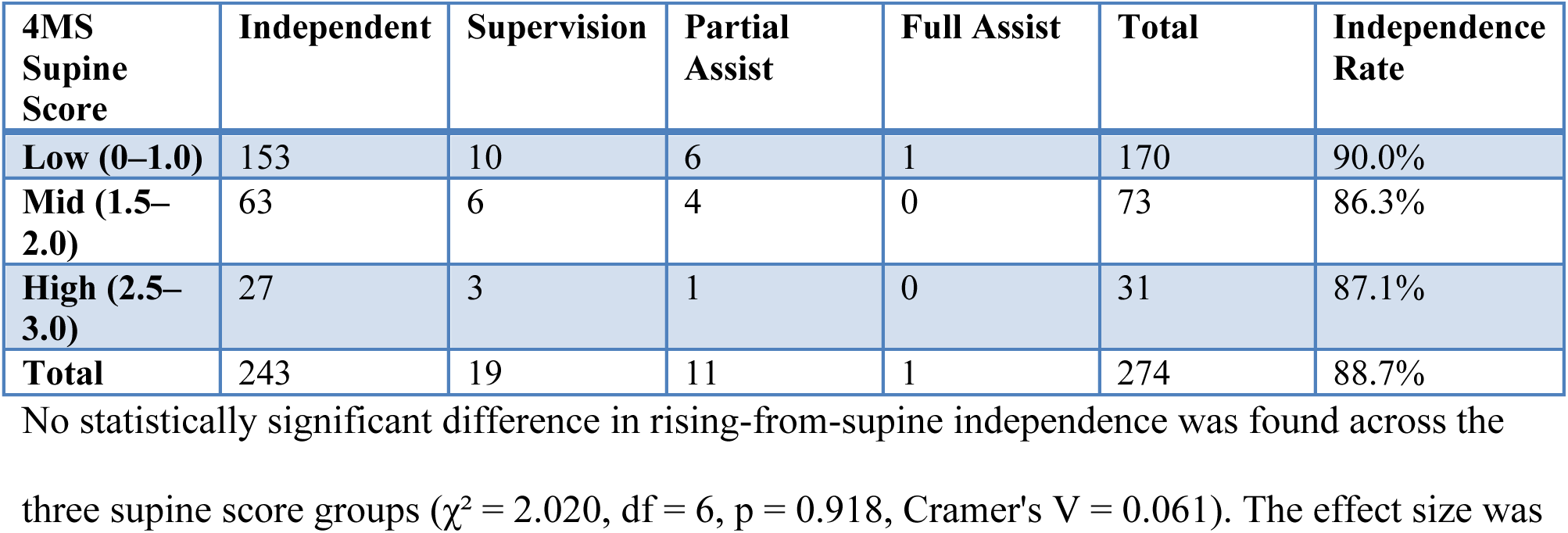
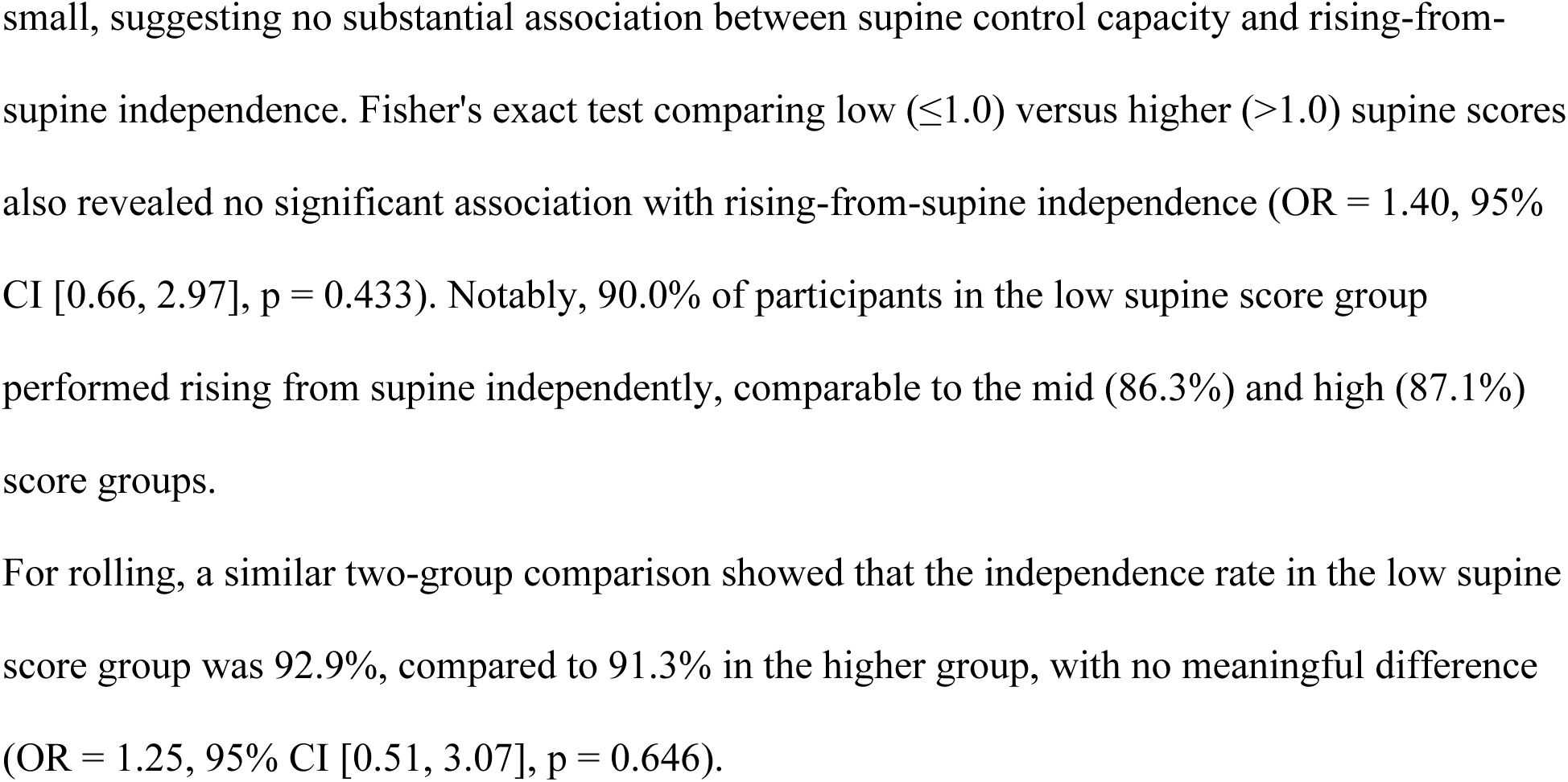
Cross-tabulation of 4MS Supine Score and Rising-from-Supine Independence (N = 274)

## Discussion

### Exploratory Findings Consistent with the 4MS Framework

The empirical results provide preliminary observations broadly consistent with the structural properties proposed by the 4MS framework. These findings generate testable hypotheses for future confirmatory research rather than constituting confirmatory evidence.

First, the moderate correlation between BI and 4MS scores (r = 0.471) is consistent with the possibility that these assessments capture different aspects of functional capacity. While BI describes task-level outcomes, 4MS is intended to reflect the structural postural control conditions underlying those outcomes. This observation is consistent with the theoretical proposition that understanding postural control structure may provide additional clinical information beyond task-based assessment.

Second, the observation of four of the five defined decline types is consistent with the non-continuity property proposed by the framework. Functional decline did not follow a single uniform pattern; rather, it presented with structurally diverse configurations that varied across individuals.

### The Supine Paradox

The most striking observation is that 74.4% of participants had the supine phase identified as the primary intervention target—despite the supine position being the most gravity-supported posture. We term this the “supine paradox.“

This finding is consistent with the compensatory preservation property proposed by the 4MS framework. Participants appear to maintain standing and walking function through compensatory strategies while exhibiting deficits in foundational supine control—including respiratory control, head stability, trunk flexion, and lower extremity elevation. Hodges and Richardson [22] demonstrated that feedforward contraction of the transversus abdominis precedes all limb movements, and Granacher et al. [23] showed that trunk muscle strength plays a critical role in balance and fall prevention in older adults. From a developmental kinesiology perspective, the neonatal trunk (0 months) is characterized by anterior pelvic tilt and rib flare, reflecting the absence of intra-abdominal pressure (IAP). By approximately 3 months, coordinated opposing activation of the diaphragm and pelvic floor establishes IAP, achieving sagittal pelvic stabilization [6, 7]. The 4MS supine phase 2-point criterion—lower extremity elevation while maintaining pelvic stability—functions as a clinically observable indicator of this IAP-generating mechanism. Age-related decline in supine scores may thus be interpreted as a structural regression toward the neonatal state of anterior pelvic tilt and rib flare—a re-collapse of the foundational IAP-generating mechanism.

Importantly, the supine paradox does not imply that participants were unable to perform supine-phase motor tasks. Quantitative analysis of bed mobility independence in 274 participants (Table 6) showed that 90.0% of those in the low supine score group (0–1.0 points, n = 170) performed rising from supine independently, with no significant difference compared to the mid (86.3%) and high (87.1%) score groups (χ² = 2.020, p = 0.918, Cramer’s V = 0.061; OR = 1.40, 95% CI [0.66, 2.97], p = 0.433). Rising-from-supine independence was broadly maintained regardless of 4MS supine score level. What the 4MS assessment detected was not the inability to perform these movements, but a qualitative deficit in the underlying motor control. Specifically, the majority of participants were unable to achieve the 4MS supine phase 2-point criterion: actively lifting the ipsilateral foot (with the knee in slight flexion and the hip in neutral rotation) and touching the toes with the ipsilateral hand while maintaining pelvic stability. This task requires coordinated activation of the deep abdominal stabilizers to anchor the pelvis; without adequate pelvic stabilization, the movement cannot be performed without compensatory strategies. The 4MS scoring system (0–3 scale) differentiates not merely whether a task can be completed, but the degree of compensatory strategy employed, thereby revealing foundational motor control deficits in individuals whom conventional ADL assessments classify as “independent.” This distinction is clinically significant: these individuals function in daily life through compensation, but the underlying deficit in deep stabilizer control may constitute a risk factor for future balance impairment and ADL decline.

Several alternative interpretations warrant consideration. First, the finding that 90.0% of participants with low supine scores performed rising from supine independently could be interpreted not as evidence of compensatory preservation, but as evidence that 4MS supine scores have limited criterion validity for predicting bed mobility outcomes. This interpretation cannot be excluded with the present data and highlights the need for future studies examining whether low supine scores predict clinically relevant outcomes such as falls or ADL decline over time. Second, low supine scores may partly reflect age-related reductions in flexibility (e.g., hamstring tightness, hip joint range-of-motion limitations) rather than deficits in deep stabilizer motor control per se. Future studies should incorporate objective flexibility measurements to disentangle these contributions. Third, an alternative hypothesis—that low supine scores reflect inherent task difficulty rather than genuine motor control deficits—warrants consideration.

However, the 4MS supine phase 2-point criterion (hip flexion with lower extremity elevation) corresponds to a postural control milestone acquired at approximately 3 months of age [5, 24], developmentally earlier than sitting (5–7 months), standing (9–13 months), or single-leg standing (12–18 months). Low supine scores are therefore more parsimoniously interpreted as reflecting foundational motor control impairment rather than task difficulty artifact.

Critically, the BI does not include any item directly assessing supine-phase postural control, covering sitting, standing, and single-leg phase tasks but not rolling or rising from supine. This structural gap may partially explain why supine-phase impairments have remained clinically underrecognized. The 4MS framework addresses this blind spot by explicitly including a supine control phase.

This finding is also consistent with the non-reversal hypothesis: functional decline does not simply progress from the most complex to the simplest posture. Instead, compensatory strategies may mask foundational impairments, creating a pattern where higher-level functions are maintained while lower-level control deteriorates.

### Asymmetry and Non-Continuity

Bilateral scores were recorded for supine, sitting, and single-leg phases. Left-right asymmetry (defined as any difference between bilateral scores) was present in 32.7% of participants (97/297) across at least one phase. Phase-specific prevalence was: supine 19.9%, sitting 12.5%, and single-leg 13.1%. Wilcoxon signed-rank tests confirmed no systematic laterality bias in any phase (all p > 0.25). These observations suggest that asymmetry may be a measurable structural property of postural control decline, consistent with the inclusion of bilateral assessment in the 4MS framework. The existence of multiple decline types—each with a distinct phase-dominant pattern—is further consistent with the proposed non-continuity property, as decline patterns varied independently across postural phases.

### Relationship to Existing Literature

The moderate correlation with BI (r = 0.471) is comparable to correlations reported between BI and other postural assessment tools such as the Berg Balance Scale [25], the Timed Up and Go test [26], and the Tinetti Performance-Oriented Mobility Assessment [27], suggesting that 4MS captures a dimension of functional capacity consistent with established measures. The finding that bed mobility independence was broadly maintained even with low supine scores is also consistent with Alexander et al.’s [28] observation that bed mobility tasks engage compensatory movement strategies in older adults. Dynamic systems theory [29] emphasizes that motor behavior emerges from the interaction of multiple subsystems, which aligns with the 4MS finding that functional decline does not follow a simple reversal of the developmental sequence.

### Clinical and Theoretical Implications

These results suggest that the 4MS framework may offer:

Complementary assessment information beyond conventional ADL scores, identifying postural control impairments that are not visible from task-based evaluation alone.

Structured clinical reasoning by providing a shared framework for explaining why ADL limitations occur, rather than simply documenting whether they exist.

Targeted intervention planning by identifying the specific postural phase requiring priority intervention, enabling individualized training programs.

The target phase algorithm used in this study identifies the postural control phase with the greatest relative deficit as the priority intervention target, with ties resolved by prioritizing developmentally earlier (lower-order) phases. When left-right asymmetry is present, the lower bilateral score determines the phase’s priority ranking. This algorithmic approach, based on the developmental sequence underlying the 4MS framework, provides a reproducible decision rule that translates evaluation data into specific training recommendations, reducing reliance on subjective clinical judgment. The finding that supine was the most frequent target phase (74.4%) underscores the prevalence of foundational postural control deficits in this population—deficits that would not be detected by conventional ADL-based assessment.

Early detection of subclinical impairments through identification of foundational postural control deficits before they manifest as overt ADL limitations.

### Limitations

This study is exploratory and hypothesis-generating in nature, and the following limitations should be acknowledged:

#### Nature of the study design

This is an exploratory cross-sectional study and is not intended to provide confirmatory validation of the structural properties proposed by the 4MS framework. The findings should be regarded as hypotheses requiring future testing through longitudinal, interventional, and multi-site replication studies.

#### Causal and temporal inference

The cross-sectional design precludes causal inference between postural control structure and ADL outcomes. Temporal ordering cannot be evaluated, and the structural properties proposed by the 4MS framework (non-continuity, asymmetry, and compensatory preservation of the non-reversal hypothesis) cannot be directly tested with cross-sectional data; confirmation of these properties requires longitudinal data.

#### Statistical interpretation

All analyses are exploratory and no formal hypothesis tests or corrections for multiple comparisons were applied. Reported p-values should be interpreted descriptively; individual significance tests should not be interpreted as confirmatory evidence.

#### Population specificity

Participants were community-dwelling day-service users with relatively high ADL independence (mean BI = 89.0). Generalization to populations with more severe functional limitations requires further investigation.

#### Single-region data

All participants were from Aomori Prefecture, Japan. Multi-site, multi-regional, and multi-national replication is needed.

#### Unestablished measurement properties

Inter-rater reliability, test-retest reliability, construct validity, and predictive validity of the 4MS evaluation have not been formally assessed. Future psychometric studies are needed to establish these properties.

#### BI ceiling effect

36.0% of participants scored the maximum BI of 100, and 55.6% scored ≥ 95. This ceiling compression limits the discriminative range of BI and likely attenuates the observed correlation. The Spearman sensitivity analysis (ρ = 0.451 vs. r = 0.471) is consistent with this interpretation. Future studies should consider using the modified BI [21] or other ADL measures with finer gradation at the upper end of the functional spectrum.

**No longitudinal follow-up to assess predictive validity.**

### Future Directions

Future research should address:

**Longitudinal studies examining the predictive validity of 4MS decline types for ADL trajectory over time.**

**Psychometric studies establishing the reliability and validity of the 4MS evaluation. Intervention studies comparing outcomes of 4MS-targeted exercise programs versus conventional approaches.**

**Multi-site and multi-national replication to establish generalizability. Integration with technology-based assessment for automated structural analysis.**

**Further investigation of the correspondence between bed mobility and 4MS scores in the supine phase. The present study provided the first quantitative description of the association between rising-from-supine/rolling independence levels and 4MS supine scores (see Supine Score and Bed Mobility Independence section above); however, detailed examination incorporating the use of environmental compensatory devices (bed rails, grab bars, electric beds) and qualitative classification of rising strategies is warranted.**

## Conclusions

The Four Movement Screen Structure (4MS) is a theoretical framework that conceptualizes the postural control structures underlying Activities of Daily Living through four distinct phases: supine, sitting, standing, and single-leg standing. This paper has presented both the theoretical basis and exploratory, preliminary empirical observations relevant to the framework.

Analysis of 297 certified care recipients revealed: (a) a moderate correlation between BI and 4MS scores (r = 0.471), consistent with the possibility that these assessments capture structurally distinct dimensions; (b) distinct patterns of functional decline, consistent with the proposed non-continuity property; (c) the “supine paradox,” in which foundational supine control was among the most commonly impaired phases despite being the most gravity-supported position, consistent with the proposed compensatory preservation property; and (d) in a subsample of 274 participants, a quantitative pattern in which 90.0% of those with low supine scores (0–1.0, n = 170) performed rising from supine independently (χ² = 2.020, p = 0.918), with no meaningful difference in rolling independence between groups (low 92.9% vs. higher 91.3%, OR = 1.25, p = 0.646), suggesting that the supine paradox may reflect qualitative motor control deficits rather than functional inability.

These exploratory and preliminary cross-sectional findings are broadly consistent with the non-reversal hypothesis proposed by the 4MS framework and suggest that 4MS may offer a complementary structural perspective for understanding ADL limitations in rehabilitation and long-term care settings. However, as this study is an exploratory, hypothesis-generating cross-sectional investigation, it does not directly establish causal relationships or temporal ordering between postural control structure and ADL outcomes, and it does not provide confirmatory evidence. The framework generates testable hypotheses for future longitudinal and confirmatory research. Future research should address psychometric investigation, longitudinal prediction studies, multi-site replication, and intervention studies to further examine the clinical utility of the framework.

**Figure 1.**
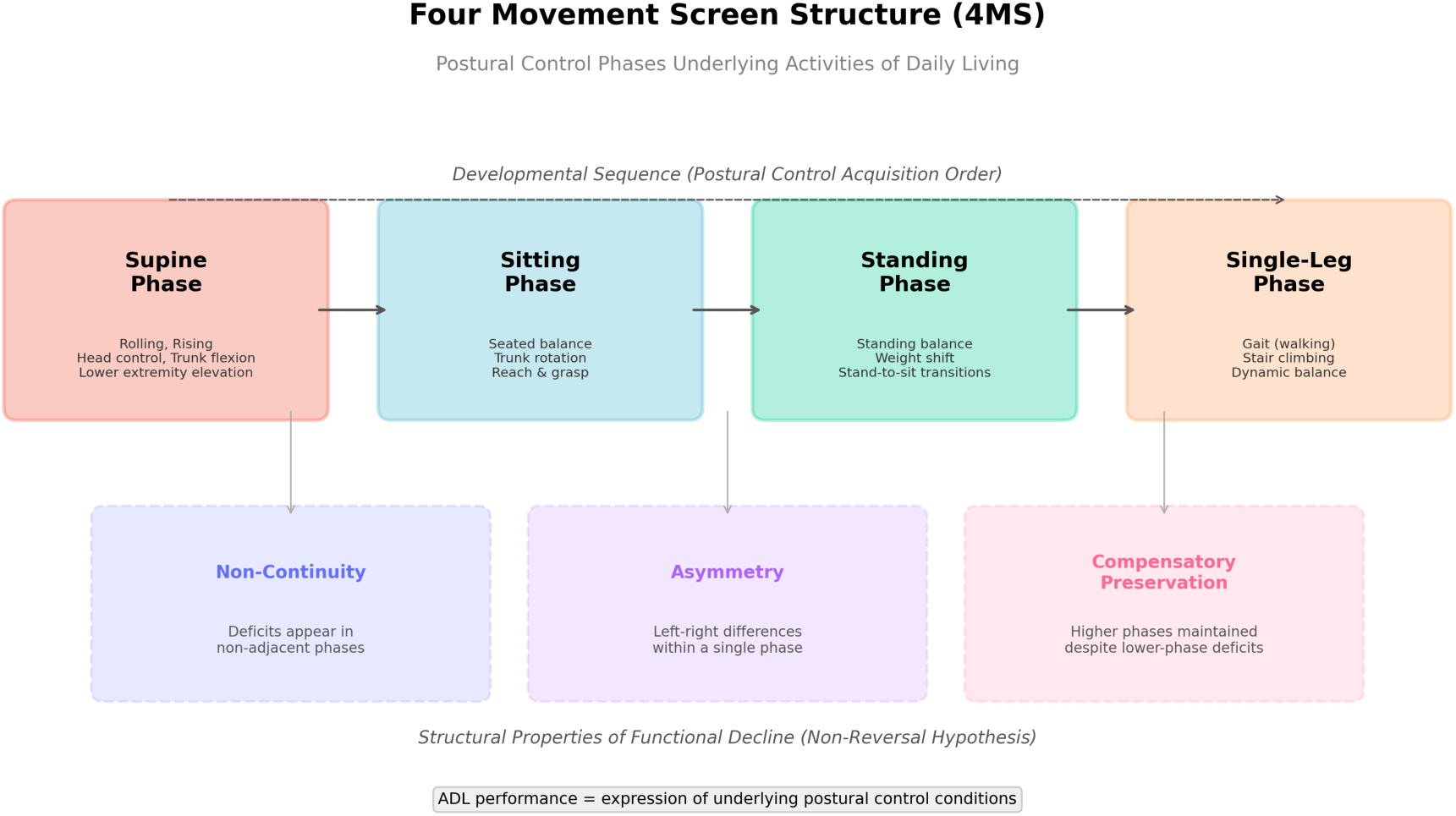
Conceptual structure of the Four Movement Screen Structure (4MS). The figure illustrates four postural control phases—supine, sitting, standing, and single-leg standing—as distinct postural control conditions underlying Activities of Daily Living (ADL). Three structural properties are indicated: Non-Continuity, Asymmetry, and Compensatory Preservation.

**Figure 2.**
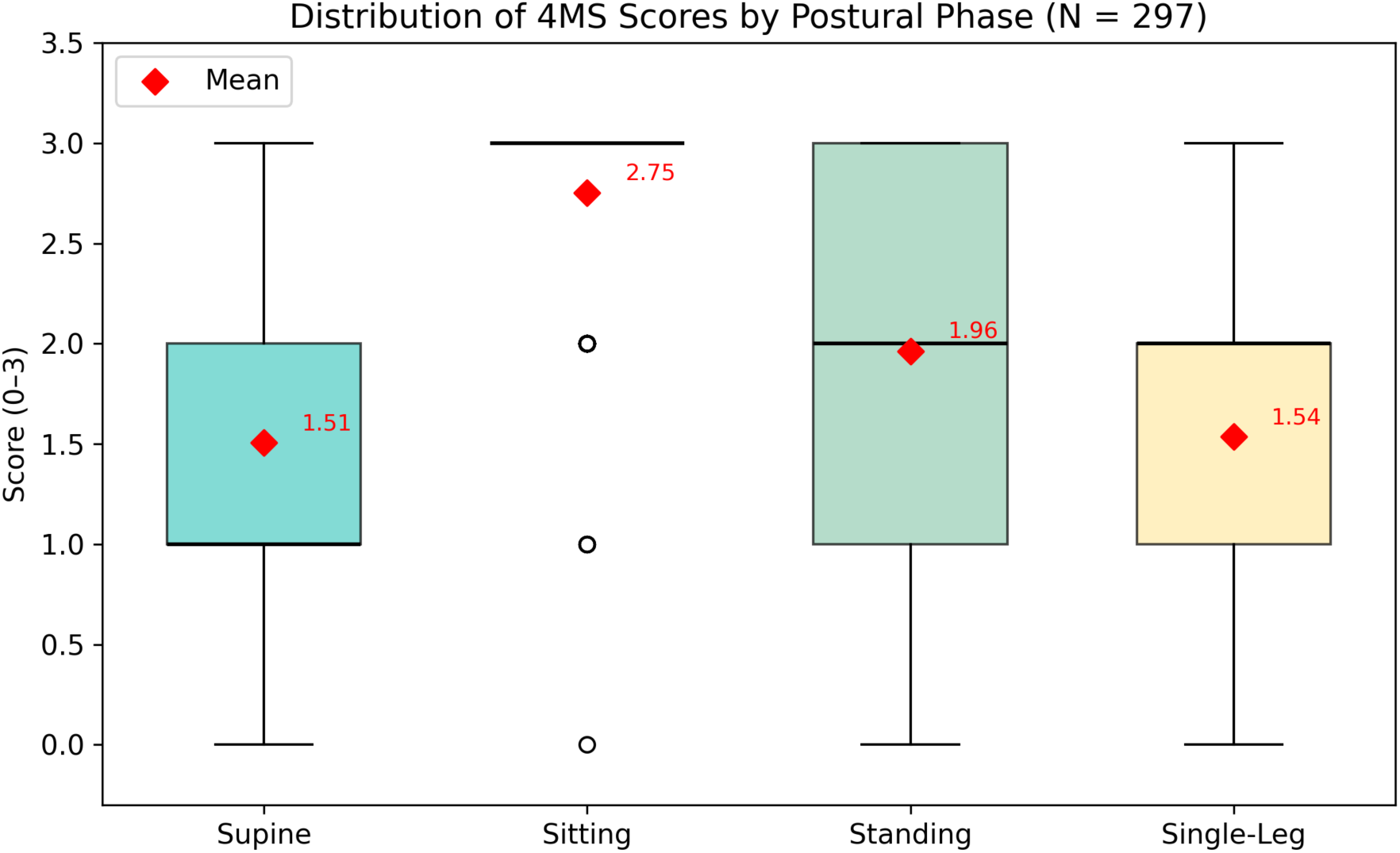
Distribution of 4MS postural control scores across four phases (N = 297). Box plots showing the score distribution (0–3 scale) for each phase. The sitting phase demonstrates the highest median with the smallest variance; the supine phase shows the lowest mean score.

**Figure 3.**
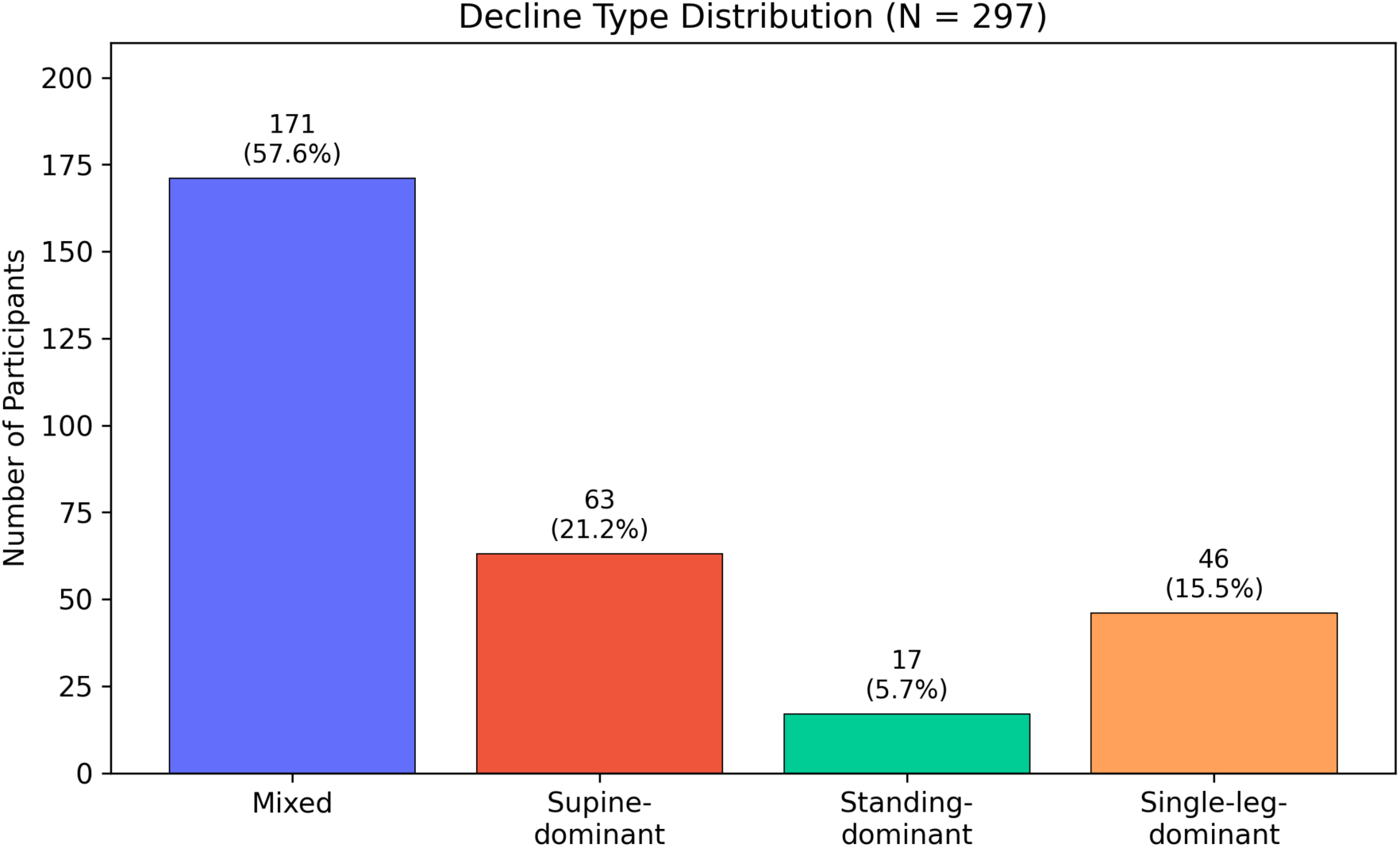
Decline type distribution in the study population. Of the five defined decline types, four were observed: mixed (57.6%), supine-dominant (21.2%), standing-dominant (5.7%), and single-leg-dominant (15.5%). Sitting-dominant was not observed.

**Figure 4.**
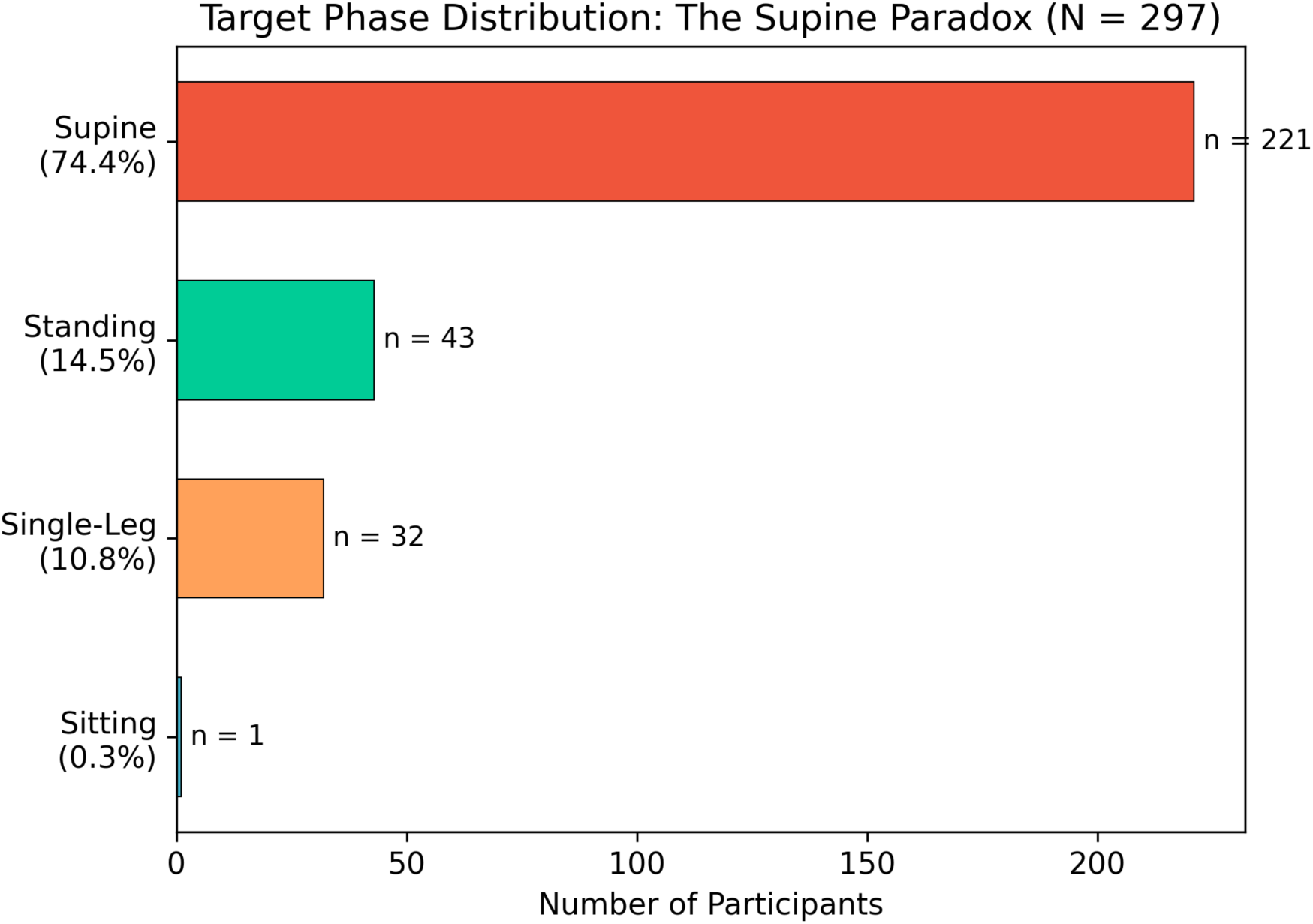
The Supine Paradox: Target phase distribution. The supine phase was identified as the primary intervention target in 74.4% of cases, despite being the most gravity-supported postural position. This finding is consistent with the compensatory preservation property proposed by the 4MS framework.

**Figure 5.**
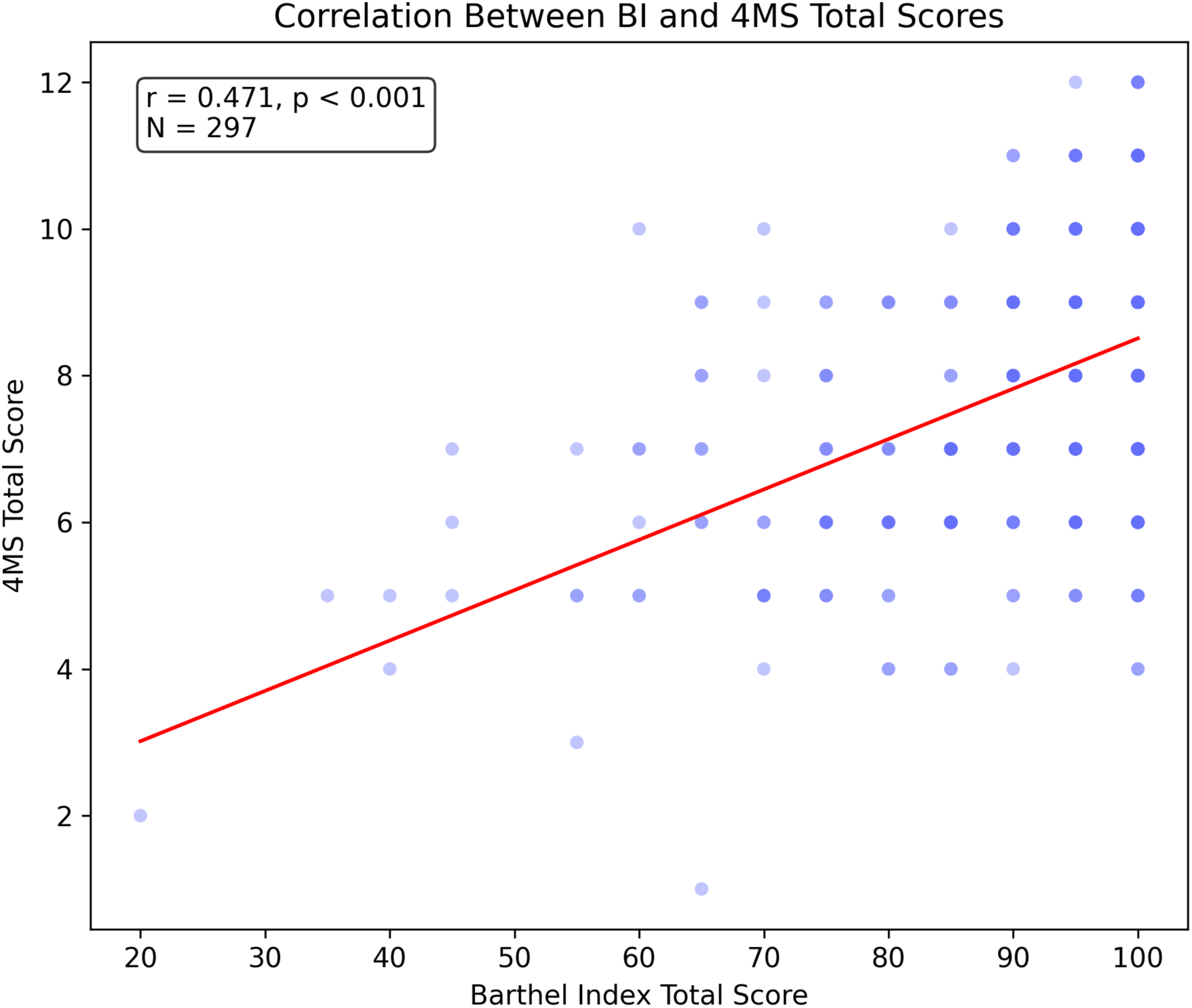
Scatter plot of BI total score versus 4MS total score (r = 0.471, N = 297). The moderate positive correlation suggests that BI and 4MS capture partially overlapping but structurally distinct aspects of functional capacity.

## Data Availability

All data produced in the present study are available upon reasonable request to the corresponding author.

## Declarations

Ethics approval and consent to participate This study utilized anonymized data collected as part of routine clinical assessment at day-service facilities operated by Care Smile Aomori Inc., under a joint research agreement with 4MS Inc. Because the study involved retrospective analysis of pre-existing anonymized clinical data with no intervention beyond standard care, formal institutional review board (IRB) approval was not required. This determination was made by the research ethics officer of Care Smile Aomori Inc. in accordance with the Japanese Ethical Guidelines for Medical and Biological Research Involving Human Subjects (Ministry of

Education, Culture, Sports, Science and Technology and Ministry of Health, Labour and Welfare, 2021 revision), which exempt retrospective observational studies using pre-existing anonymized clinical data from mandatory ethics review. All data were anonymized prior to analysis, with no personally identifiable information retained. The study was conducted in accordance with the Declaration of Helsinki. Written informed consent for the routine clinical assessments was obtained as part of standard facility procedures.

Consent for publication Not applicable. All data were anonymized.

Availability of data and materials The datasets analyzed during the current study are available from the corresponding author on reasonable request.

Competing interests Hiroshi Osato is the CEO of 4MS Inc., which developed the 4MS evaluation system. This potential conflict of interest is declared in the interest of full transparency. To mitigate potential bias, the study employed pre-defined classification rules (decline type, target phase), reported all analyses including null findings, and made the analytical approach fully transparent to enable independent replication. Data collection was performed by trained clinical staff at the participating facilities under routine care procedures, independent of the author’s direct supervision.

Funding The 4MS system development has received domestic innovation support. Related patents: Japanese Patent No. 6901809 (ADL improvement support device), with international registrations in China and Hong Kong; and Japanese Patent No. 7790790 (Physical function exercise provision processing system).

Authors’ contributions Hiroshi Osato conceived the theoretical framework, designed the study, supervised data collection, performed the analysis, and wrote the manuscript.

## Acknowledgements

The author acknowledges the clinical staff at participating facilities, and the long-term care users who participated in routine assessments. The author also thanks the Aomori Prefecture intellectual property support organizations for their guidance.

## Supplementary Materials

**Supplementary Table S1.** Distribution of IADL Performance Levels (N = 240)

| Item | Independent | Supervision | Partial assistance | Full assistance |
| --- | --- | --- | --- | --- |
| Cooking | 65 (27.1%) | 3 (1.2%) | 43 (17.9%) | 129 (53.8%) |
| Laundry | 82 (34.5%) | 3 (1.3%) | 46 (19.3%) | 107 (45.0%) |
| Cleaning | 61 (25.7%) | 7 (3.0%) | 56 (23.6%) | 113 (47.7%) |

**Supplementary Table S2.** Distribution of Basic Movement Performance Levels (N = 274)

| Item | Independent | Supervision | Partial assistance | Full assistance |
| --- | --- | --- | --- | --- |
| Rolling | 253 (92.3%) | 9 (3.3%) | 10 (3.6%) | 2 (0.7%) |
| **Rising from supine** | 243 (88.7%) | 19 (6.9%) | 11 (4.0%) | 1 (0.4%) |
| **Sitting** | 266 (97.1%) | 6 (2.2%) | 2 (0.7%) | 0 (0.0%) |
| **Standing up** | 235 (85.8%) | 31 (11.3%) | 7 (2.6%) | 1 (0.4%) |
| **Standing** | 235 (85.8%) | 28 (10.2%) | 9 (3.3%) | 1 (0.4%) |

Note: Standing data available for 273 of 274 participants (1 case missing).

## References

1. Mahoney FI, Barthel DW. Functional evaluation: the Barthel Index. Md State Med J. 1965;14:61–65.

2. Granger CV, Hamilton BB, Keith RA, Zielezny M, Sherwin FS. Advances in functional assessment for medical rehabilitation. Top Geriatr Rehabil. 1986;1(3):59–74. 10.1097/00013614-198604000-00007.

3. Keith RA, Granger CV, Hamilton BB, Sherwin FS. The Functional Independence Measure: a new tool for rehabilitation. Adv Clin Rehabil. 1987;1:6–18.

4. Hadders-Algra M. Development of postural control during the first 18 months of life. Neural Plast. 2005;12(2-3):99–108. 10.1155/np.2005.99.

5. Vojta V. Die zerebralen Bewegungsstörungen im Säuglingsalter: Frühdiagnose und Frühtherapie. Stuttgart: Enke Verlag; 1974.

6. Frank C, Kobesova A, Kolář P. Dynamic neuromuscular stabilization & sports rehabilitation. Int J Sports Phys Ther. 2013;8(1):62–73.

7. Kolář P, Bitnar P, Dyrhonová O, Horáček O, Kříž J. Clinical Rehabilitation. Prague: Rehabilitation Prague School; 2013.

8. Bly L. Motor Skills Acquisition in the First Year: An Illustrated Guide to Normal Development. Tucson, AZ: Therapy Skill Builders; 1994.

9. Gallahue DL, Ozmun JC, Goodway JD. Understanding Motor Development: Infants, Children, Adolescents, Adults. 7th ed. New York: McGraw-Hill; 2012.

10. Gesell A, Amatruda CS. Developmental Diagnosis: Normal and Abnormal Child Development. 2nd ed. New York: Paul B. Hoeber; 1947.

11. McGraw MB. The Neuromuscular Maturation of the Human Infant. New York: Columbia University Press; 1943.

12. Piper MC, Darrah J. Motor Assessment of the Developing Infant. Philadelphia: W.B. Saunders; 1994.

13. WHO Multicentre Growth Reference Study Group. WHO Motor Development Study: windows of achievement for six gross motor development milestones. Acta Paediatr. 2006;95(Suppl 450):86–95. 10.1111/j.1651-2227.2006.tb02379.x.

14. Bernstein NA. The Co-ordination and Regulation of Movements. Oxford: Pergamon Press; 1967.

15. Horak FB, Nashner LM. Central programming of postural movements: adaptation to altered support-surface configurations. J Neurophysiol. 1986;55(6):1369–1381. 10.1152/jn.1986.55.6.1369.

16. Horak FB. Postural orientation and equilibrium: what do we need to know about neural control of balance to prevent falls? Age Ageing. 2006;35(S2):ii7–ii11. 10.1093/ageing/afl077.

17. Massion J. Postural control system. Curr Opin Neurobiol. 1994;4(6):877–887. 10.1016/0959-4388(94)90137-6.

18. Reisberg B, Kenowsky S, Franssen EH, et al. Towards a science of Alzheimer’s disease management: a model based upon current knowledge of retrogenesis. Int Psychogeriatr. 1999;11(1):7–23. 10.1017/s1041610299005554.

19. Shumway-Cook A, Woollacott MH. Motor Control: Translating Research into Clinical Practice. 5th ed. Philadelphia: Wolters Kluwer; 2017.

20. Collin C, Wade DT, Davies S, Horne V. The Barthel ADL Index: a reliability study. Int Disabil Stud. 1988;10(2):61–63. 10.3109/09638288809164103.

21. Shah S, Vanclay F, Cooper B. Improving the sensitivity of the Barthel Index for stroke rehabilitation. J Clin Epidemiol. 1989;42(8):703–709. 10.1016/0895-4356(89)90065-6.

22. Hodges PW, Richardson CA. Feedforward contraction of transversus abdominis is not influenced by the direction of arm movement. Exp Brain Res. 1997;114(2):362–370. 10.1007/pl00005644.

23. Granacher U, Gollhofer A, Hortobagyi T, Kressig RW, Muehlbauer T. The importance of trunk muscle strength for balance, functional performance, and fall prevention in seniors: a systematic review. Sports Med. 2013;43(7):627–641. 10.1007/s40279-013-0041-1.

24. Hadders-Algra M. Typical and atypical development of reaching and postural control in infancy. Dev Med Child Neurol. 2013;55(Suppl 4):5–8. 10.1111/dmcn.12298.

25. Berg KO, Wood-Dauphinee SL, Williams JI, Maki B. Measuring balance in the elderly: validation of an instrument. Can J Public Health. 1992;83(Suppl 2):S7–S11.

26. Podsiadlo D, Richardson S. The timed “Up & Go“: a test of basic functional mobility for frail elderly persons. J Am Geriatr Soc. 1991;39(2):142–148. 10.1111/j.1532-5415.1991.tb01616.x.

27. Tinetti ME. Performance-oriented assessment of mobility problems in elderly patients. J Am Geriatr Soc. 1986;34(2):119–126. 10.1111/j.1532-5415.1986.tb05480.x.

28. Alexander NB, Fry-Welch DK, Ward ME, Folkmier LC. Bed mobility task performance in older adults. J Rehabil Res Dev. 2000;37(5):633–638.

29. Thelen E, Smith LB. A Dynamic Systems Approach to the Development of Cognition and Action. Cambridge, MA: MIT Press; 1994.

